# Assessing PMMoV as a faecal marker for wastewater-based surveillance - Insights from Swedish wastewaters and foods

**DOI:** 10.1101/2025.05.18.25327821

**Authors:** Sofia Persson, Javier Edo Varg, Sarah Coker, Fanny Persson, Filip Petrini, Israa Dafalla, Lauren Davies, Polina Vaulina, Monika Vilemova, Nahla Mohamed, Maja Malmberg, Anna J. Székely

## Abstract

Pepper Mild Mottle Virus (PMMoV) is widely used in wastewater-based surveillance (WBS) as a faecal indicator, population size marker and analytical process control. It is highly abundant in domestic wastewater and originates from consumption of products containing *Capsicum* species infected with PMMoV. However, despite its widespread use in WBS, data on the variability of PMMoV levels in wastewater are limited, and the main dietary sources are not fully investigated. In this study, we assessed the reliability of PMMoV as a faecal marker by analysing wastewater samples collected across multiple locations and timeframes in Sweden. We also analysed different foods to identify possible dietary sources. Our results showed consistently high PMMoV levels in wastewater, averaging 10.37 log_10_ genome copies per resident per day, with overall little variation between days, weeks, and years. Of 60 foods tested, 45 contained detectable levels of PMMoV, with particularly high concentrations, ranging from 5.70 to 12.21 log_10_ genome copies per serving, found in commonly consumed spices, including paprika powder, and various processed foods, including ready meals, sandwich spreads and snack products. These results highlight the dietary basis for the high and stable presence of PMMoV in domestic wastewater and emphasise its robustness as a faecal marker in the Swedish context.

## Introduction

Wastewater-based surveillance (WBS) is a powerful tool for monitoring infectious viral diseases in communities, serving as a valuable complement to clinical testing (1,2). To date, WBS has been successfully implemented in over 70 countries (3), enabling the monitoring of various viral pathogens, including hepatitis A virus (4), influenza A and B viruses (5,6), noroviruses (7), poliovirus (8), respiratory syncytial virus and SARS-CoV-2 (9). However, fluctuations in virus levels in wastewater do not always correspond to changes in the number of infected individuals. These variations can also result from changes in wastewater flow due to factors such as snowmelt and precipitation, fluctuations in the number of individuals contributing to the system, environmental conditions like pH and temperature, as well as differences in sampling and analytical procedures (1,2,10).

To account for these sources of variability and improve data comparability over time and across wastewater treatment plants (WWTPs), normalisation strategies are essential. One common normaliser is the use of a human faecal marker, which estimates the size of the contributing population and serves as a process control during analysis by accounting for changes in the wastewater matrix that may affect target virus stability (1). A good human faecal marker should be highly abundant in human faeces, consistently excreted across different time periods and geographical locations, and exhibit similar behaviour in wastewater as the target pathogen(s) (1,7,11).

One of the most commonly used faecal markers is Pepper Mild Mottle Virus (PMMoV), a non-enveloped, single-stranded RNA virus and one of the most abundant RNA viruses in human faeces (12–15). PMMoV infects *Capsicum* plants, such as peppers and chillies, and is introduced to humans through dietary consumption (15,16). Importantly, PMMoV is rarely detected in animal faecal samples, underlining its specificity as a marker for human faecal contamination (14,16,17). It is both highly abundant and highly stable in domestic wastewater, with concentrations of 5-10 log_10_ genome copies (gc)/l in untreated wastewater, making it quantifiable even in small sample volumes (7,16). In addition, PMMoV often serves as a process control for the sample concentration and nucleic acid extraction process, as it is analysed together with the pathogenic viruses of interest.

Despite its many advantages and widespread use in WBS, the reliability of PMMoV as a faecal marker has been questioned. Several studies have reported that alternative normalisation methods, or even the use of non-normalised data, correlate more strongly with clinical case data (18–20). Investigations into the spatiotemporal variability of PMMoV in wastewater has been conducted in regions such as North America (10,11,21,22), Germany (23) and Saudi Arabia (24). However, there is a lack of detailed studies from the Nordic countries. Given that factors such as climate, infrastructure and behavioural and dietary patterns may influence the effectiveness of faecal markers, site-specific validation is essential to ensure their reliability across diverse settings.

A commonly debated factor contributing to variability in PMMoV levels is the consumption of *Capsicum*-containing foods, which can fluctuate over time and among populations with differing dietary habits (25). Previous research has speculated that variations in PMMoV concentrations in wastewater may be linked to dietary differences, particularly the intake of pepper-based products such as hot sauces, spices and salsa (10), but no follow-up studies have been conducted to verify these associations. Existing studies on PMMoV in food have primarily focused on spicy items such as gochujang, chilli sauces, red chilli peppers and cayenne pepper (15,26,27). Nevertheless, PMMoV-rich foods are not necessarily restricted to spicy products, and the presence of the virus in a broader range of foods remains largely unexplored.

On the one hand, it can be hypothesised that PMMoV may be a poor faecal marker, as demographic and socioeconomic differences, such as the proportion of foreign-born residents or overall socioeconomic status, could influence *Capsicum* consumption, leading to variations in PMMoV excretion. Dietary patterns, strongly shaped by cultural background, often remain stable over a person’s lifetime (28). Traditionally, *Capsicum*-rich cuisines are more prevalent in warmer regions such as South America, Asia and Africa, whereas colder regions like Sweden have historically favoured milder flavours.

On the other hand, globalisation has increased the availability and popularity of *Capsicum*-containing foods, such as Thai, Indian, and Mexican cuisine, potentially reducing dietary differences between populations. Processed foods flavoured with chilli or paprika are also widely consumed. As a result, variations in *Capsicum* intake may be less pronounced than previously assumed. However, certain foods, like chilli-flavoured snacks or tacos, may be eaten more often on specific days; for instance, “Taco Friday” is a well-known tradition in Sweden (29). Such patterns could contribute to short-term fluctuations in PMMoV excretion and daily variability in wastewater concentrations.

In this study, we assess the suitability of PMMoV as a faecal indicator and population size marker in Sweden. First, we examined its spatiotemporal variability across 18 WWTPs representing diverse community sizes, socioeconomic profiles and geographical locations, over a six-month period. Second, we analysed long-term temporal variability and seasonal variability through weekly sampling over a 34-month period at four geographically distinct WWTPs. Third, we investigated short-term fluctuations by conducting daily sampling at a single WWTP over a period of five weeks. Finally, to identify potential dietary sources of PMMoV, we analysed 60 different food products of different categories, focusing mainly on those containing *Capsicum*-derived ingredients.

## Methods

### Wastewater samples

PMMoV sampling and analysis in raw wastewater were conducted as part of the monitoring of human respiratory viruses by the Swedish Environmental Epidemiology Centre (SEEC) at the Swedish University of Agricultural Sciences (SLU) (30).

#### Sampling sites and sampling period

This study included three sampling series: short-term, long-term and daily monitoring (Table 1). The short-term monitoring aimed to investigate potential regional variations in PMMoV levels and examine the relationship between total daily PMMoV load and the number of residents connected to each WWTP. The long-term monitoring focused on assessing temporal variations in PMMoV levels over an extended period, while the daily monitoring aimed to evaluate day-to-day fluctuations in PMMoV levels. The number of residents connected to each WWTP was provided by the respective WWTPs (Table 1).

**Table 1.** Sample collection dates and number of samples collected at each site. Number of samples included denote samples with valid quality control characteristics. An asterisk (*) next to the number of residents indicates that the value is an estimate of the people connected based on a biological oxygen demand (BOD-7) value, rather than the number of residents physically connected to the WWTP

| City | Monitoring | Number of residents connected to the WWTP | Sample collection, start date | Sample collection, end date | Number of samples analysed | Number of samples included |
| --- | --- | --- | --- | --- | --- | --- |
| Gävle | Short-term | 89,000 | 01-01-2024 | 07-01-2024 | 26 | 26 |
| Göteborg | Short-term | 830,000 | 01-01-2024 | 07-01-2024 | 27 | 26 |
| Helsingborg | Short-term | 149,000 | 01-01-2024 | 07-01-2024 | 27 | 27 |
| Kalmar | Short-term, long term | 66,600 | 05-30-2022 | 04-21-2025 | 142 | 141 |
| Karlstad | Short-term | 70,000 | 01-01-2024 | 07-01-2024 | 27 | 23 |
| Linköping | Short-term | 154,000 | 01-01-2024 | 07-01-2024 | 27 | 27 |
| Luleå | Short-term | 70,000 | 01-01-2024 | 07-01-2024 | 26 | 26 |
| Malmö | Short-term | 356,000 | 01-01-2024 | 07-01-2024 | 27 | 27 |
| Örebro | Short-term | 137,000 | 05-30-2022 | 04-14-2025 | 149 | 146 |
| Östersund | Short-term | 54,000 | 01-01-2024 | 07-01-2024 | 27 | 27 |
| Östhammar | Short-term | 4,500* | 01-01-2024 | 07-01-2024 | 26 | 26 |
| Stockholm-Bromma | Short-term | 369,000 | 01-01-2024 | 07-01-2024 | 27 | 27 |
| Stockholm-Grödinge | Short-term | 346,500 | 01-01-2024 | 07-01-2024 | 27 | 27 |
| Stockholm-Henriksdal | Short-term | 861,000 | 01-08-2024 | 07-01-2024 | 25 | 25 |
| Stockholm-Käppala | Short-term | 507,600* | 01-01-2024 | 07-01-2024 | 25 | 25 |
| Umeå | Short-term,<br>long-term | 109,300 | 05-30-2022 | 04-21-2025 | 144 | 135 |
| Uppsala | Short-term,<br>long-term,<br>daily | 198,000 | 05-30-2022 | 04-14-2025 | 182 | 179 |
| Västerås | Short-term | 145,000 | 01-08-2024 | 07-01-2024 | 26 | 26 |

For the short-term monitoring, a total of 477 samples of untreated wastewater were collected weekly from 18 WWTPs between January 2024 and July 2025. These samples represented approximately 4,516,000 residents, accounting for 42% of the Swedish population (31). For the long-term monitoring, 582 samples were collected weekly from four WWTPs between May 2022 and April 2025, covering approximately 511,000 residents or 4.72% of the Swedish population (31). During the daily monitoring, 35 samples were collected from a WWTP in Uppsala over five weeks, from 7 January to 10 February 2025. Figure 1 shows the location of the different WWTPs. Holidays, sampler errors and logistics issues resulted in the absence of some samples, leading to minor gaps in data coverage.

**Figure 1.**
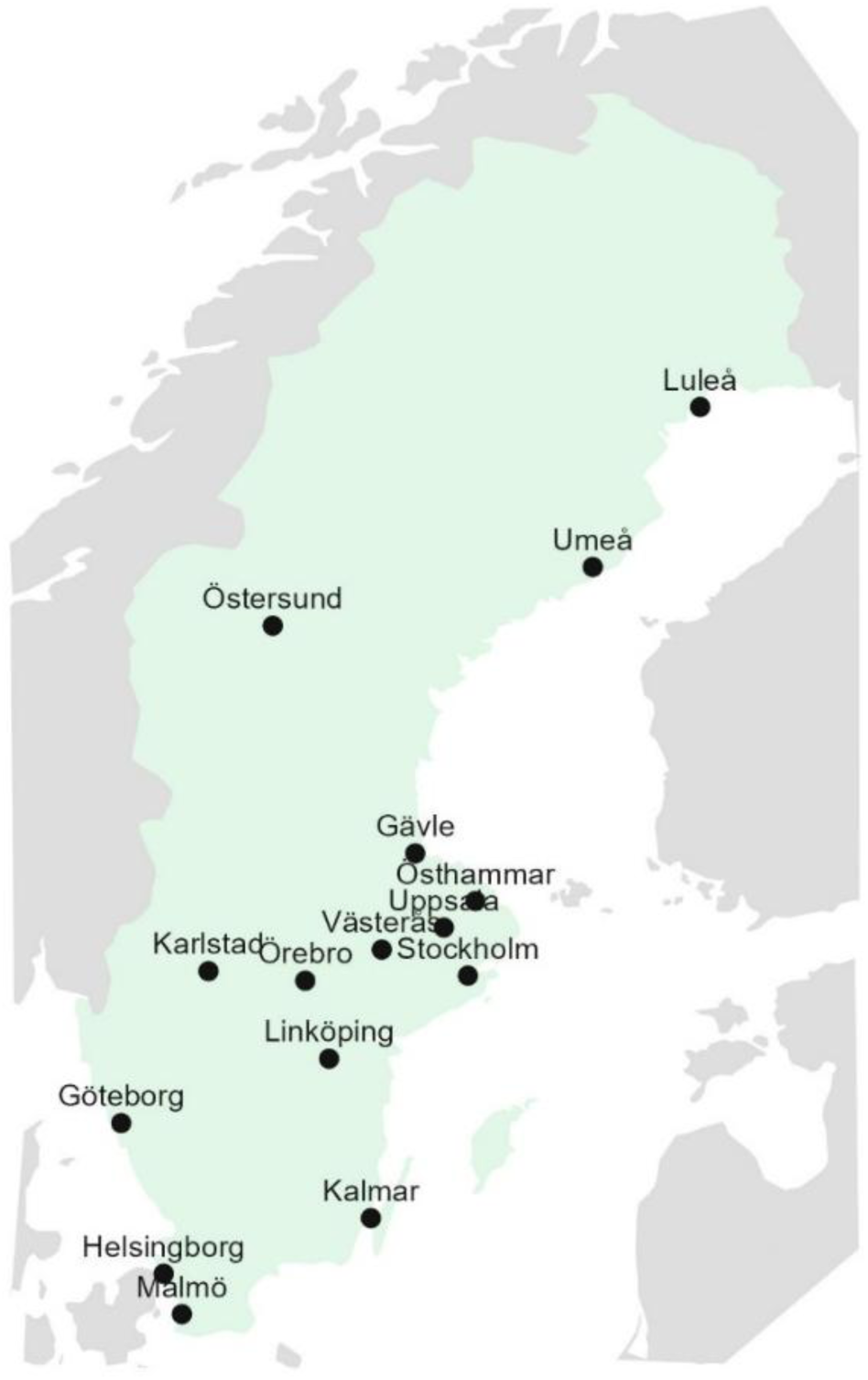
A map of Sweden, indicating the location of the WWTPs included in the study

#### Sampling, transport and collection of flow and temperature data

In most WWTPs, raw, untreated wastewater samples representative of a single day were collected with flow-compensated auto samplers. For Stockholm-Bromma, Stockholm-Henriksdal and Uppsala, samples were collected from several inlets and then flow-proportionally mixed into one sample, as described previously [(32); Table S7 in the article]. In Uppsala, samples were collected every day instead of once a week and then flow-proportionally combined into a weekly composite sample. However, for the daily monitoring in Uppsala, the daily samples were analysed individually without mixing.

Upon collection, the samples were stored at 4°C and transported to the laboratory in cold packs, where they were stored at 4°C until nucleic acid extraction the next day. The total daily wastewater flow (m^3^ per day) was recorded by each WWTP on the day of sampling, and converted to litres per day. In the cases where flow values were missing or if the flow value was obviously entered in a wrong way in our data, the flow was set to the average flow of the respective WWTP. In total, 29 samples (6.08 % of the data points) lacked associated flow data in the short-term monitoring and 27 samples (4.64%) in the long-term monitoring. Details of all samples with missing flow values are provided in Supplementary Table 1.

For WWTPs involved in long-term monitoring (Kalmar, Örebro, Umeå and Uppsala), daily average wastewater temperatures were provided by the respective WWTPs during 2024 to investigate potential associations between PMMoV levels and wastewater temperature.

#### Nucleic acid extraction

Total nucleic acids were extracted from raw wastewater using the Maxwell® RSC Enviro TNA kit (Promega, AS1831) according to the manufacturer’s instructions using the Maxwell® RSC instrument (Promega, AS4500) with the PureFood GMO and Authentication programme, as described previously (30). Each wastewater sample was divided into two subsamples. Forty ml of each subsample was used as input and the subsamples were eluted in 100 µl of nuclease-free water. A negative extraction control consisting of 40 ml of tap water or ultrapure Milli-Q water (VWR) was included in each extraction round. All nucleic acid extracts were immediately stored at -80°C until RT-qPCR the next day.

#### Collection of demographic and socioeconomic data

To investigate the relationship between demographic and socioeconomic factors and PMMoV levels in wastewater, we compiled and curated the latest open-access data (CC0 license) from Statistics Sweden’s Statistical Database (33) for each municipality served by the WWTPs included in the short-term monitoring. The dataset included municipal classifications (commuter, larger, or metropolitan city), the proportion of foreign-born residents (2023), and a socioeconomic index (2022).

The socioeconomic index represents the average of three indicators: the percentage of residents with only pre-secondary education, the percentage with a low economic standard, and the percentage either receiving economic assistance or experiencing long-term unemployment. Higher index values indicate greater socioeconomic vulnerability.

For Stockholm WWTPs serving multiple municipalities, we calculated population-weighted averages to generate a composite estimate for each plant. However, it is important to note that municipal boundaries often extend beyond the actual catchment areas, meaning the demographic data may not precisely align with the WWTP service areas.

### Food samples

#### Sample collection

To explore potential dietary sources of PMMoV in human wastewater, we collected 60 different food samples of different categories, such as spices and spice mixes, ready-made sauces, various processed foods including snacks, spreads, bouillon cubes, fresh and frozen ready-made dishes, as well as different *Capsicum* fruits. Most foods were known to contain *Capsicum* species, however, some were included without prior knowledge of their *Capsicum* content. Beyond food products, we also tested a tobacco sample (Snus) as a potential alternative source of PMMoV, since the virus has been reported to infect *Nicotiana* plants (34). In addition, oats were included as a negative control to represent a food unlikely to contain PMMoV. Most samples were purchased from local supermarkets. Detailed descriptions of the samples and their origin are presented in Supplementary Table 2.

#### Sample processing and nucleic acid extraction

Ribonucleic acid from food samples was extracted using the Maxwell® RSC Plant RNA Kit (Promega, AS1500) using the Maxwell® RSC instrument (Promega, AS4500) with the Plant RNA programme. The majority of the food samples were extracted in two subsamples.

For all samples except for the *Capsicum* fruits, a portion of the sample material was collected and homogenised using one of the following methods: vortex mixing, grinding with a mortar and pestle, or fine chopping with two sterile scalpels (the processing method of each sample is specified in Supplementary Table 2). Then, 20-200 mg of homogenate was collected and transferred directly by an inoculation loop to a bead beating tube (PowerBead Pro Tubes, Ceramic 1.4 mm from Qiagen, 13113-50) on a scale. 600 μl of chilled 1-thioglycerol/homogenising solution (supplemented with the RNA extraction kit) was added upon weighing the sample, whereupon bead beating (Bead Mill 24 device, fisherbrand™, 15515799) was performed for 5 min.

Most samples represented homogeneous portions of the tested items. However, due to the high complexity, for certain samples only specific parts of the food were analysed. These included, for instance, the filling of ready-made tuna and club sandwiches, the surface toppings of ready-made pan pizzas and various components of other ready-made foods. These samples are indicated in Supplementary Table 2.

For *Capsicum* fruits, different parts of the fruit were excised with a scalpel, blended and ground to a fine powder in liquid nitrogen using a sterile mortar and pestle. The powder was transferred to a Falcon tube and immediately placed on dry ice. One hundred mg of the powder was then transferred to a bead beating tube (PowerBead Pro Tubes, Ceramic 1.4 mm from Qiagen, 13113-50) and placed on dry ice again. Then, 600 μl of chilled 1-thioglycerol/homogenising solution was added to each subsample and bead beating was performed in pulses of 30 s, followed by deposition on wet ice until the samples were processed further.

For all sample types, the procedure was then continued according to the kit manufacturer’s instructions. The subsamples were eluted in 50-100 µl of nuclease-free water. At least one negative extraction control consisting of 100 µl of nuclease-free water was included in each extraction round, and processed in the same way as the samples. All nucleic acid extracts were immediately stored at -80°C until RT-qPCR.

### RT-qPCR

RT-qPCR was performed on a CFX duet or a CFX 96 real-time PCR system (Bio-Rad) using the Reliance One-Step Multiplex Supermix Kit (Bio-Rad; art. nr. 12010221) with 1 mg/ml bovine serum albumin (Thermo Fisher, cat. nr. AM2618). Each reaction contained 15 µl of master mix and 5.0 µl of template, with 600 nM forward primer, 800 nM reverse primer and 200 nM probe (Table 2).

**Table 2.**
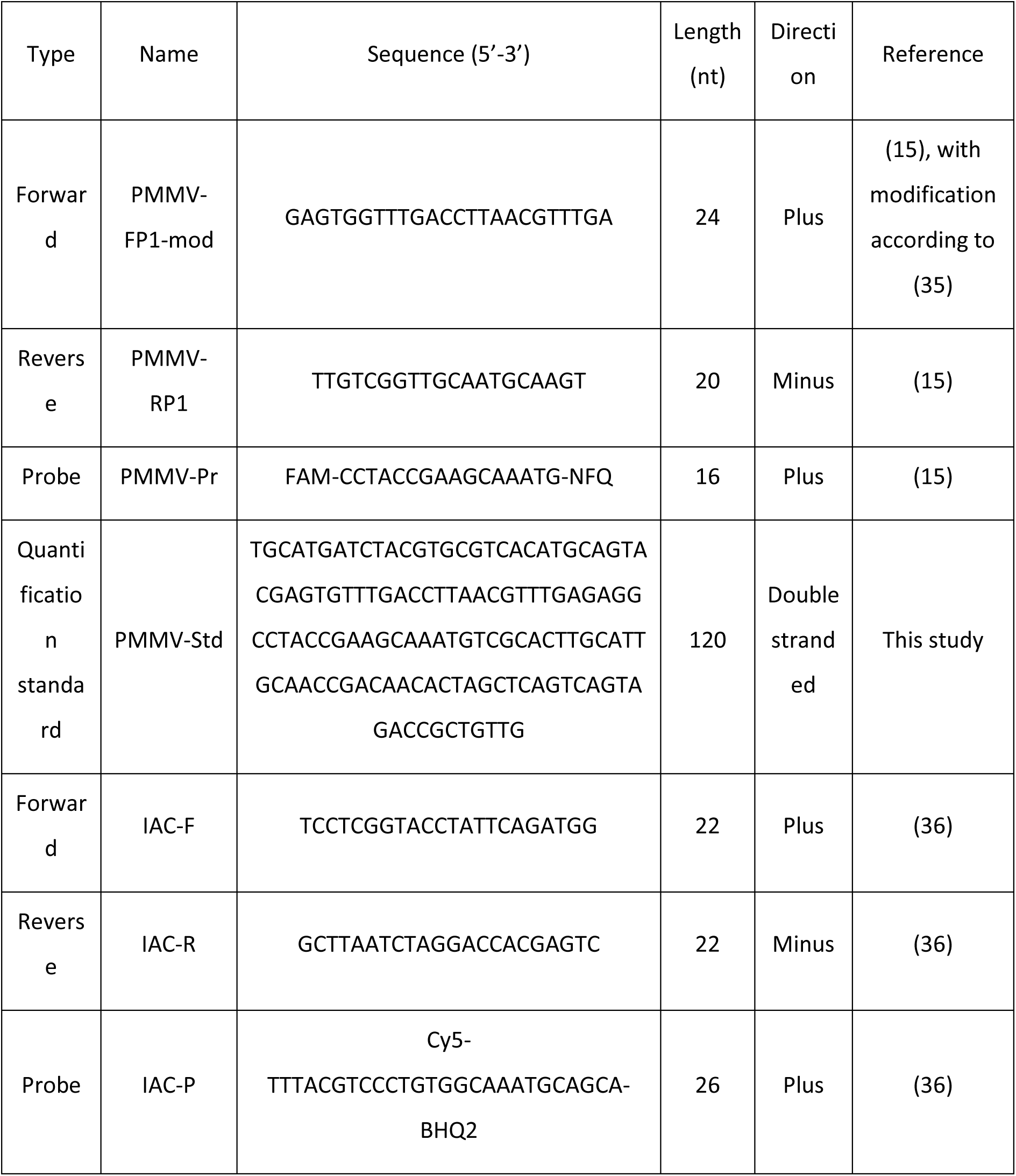
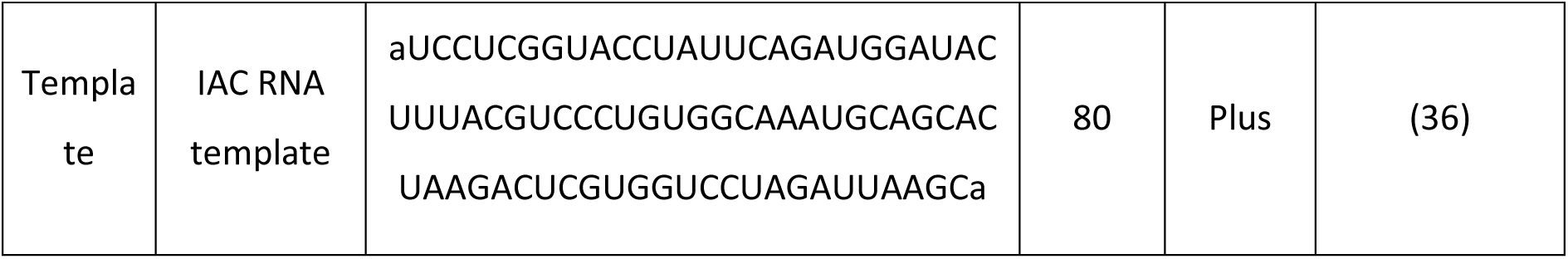
Primers, probes, quantification standards and internal controls

Starting from week 12, 2024 for the wastewater samples and for all food samples, an internal amplification control (IAC) was incorporated to assess RT-qPCR inhibition. The IAC reactions included 400 nM forward primer, 400 nM reverse primer, 200 nM probe, and 10^4^ copies of synthetic RNA template (Table 2), and reactions were performed in duplex format. The primers, probe and synthetic RNA template were purchased from Intergrated DNA Technologies (IDT).

The thermal profile was as follows: 50°C for 30 min and 95°C for 5 min, followed by 45 cycles of 95°C for 15 s and 58°C for 30 s. Results were analysed using CFX Maestro Software version 2.3 (Bio-Rad) and the cycle of quantification (Cq) values were determined using the regression method within the analysis software.

Each plate contained two wells of each subsample (extracted sample), negative extraction controls (NECs), no-template controls (NTCs) and, starting in week 35 2022 for the wastewater samples, positive controls consisting of archived wastewater TNA extracts positive for PMMoV. A gBlock standard (Table 2) was used for quantification. The standard was run as a tenfold dilution series ranging from 10^6^ to 100 copies per reaction, and with two wells per dilution level. Obtained concentrations were converted to correspond to the number of gc per llitre for wastewater samples and to the number of gc per gram for food samples, and then log_10_ transformed before further analysis. For each sample, the average value of the two RT-qPCR wells was calculated, followed by averaging across the two subsamples, yielding the final value used in subsequent analyses.

### Quality control

The quality control procedures included establishing a limit of blank, testing for inhibition (beginning in week 12, 2024 for the wastewater samples) and evaluating variability between replicate extractions (subsamples).

Inhibition in RT-qPCR was assessed by comparing the IAC Cq values. Specifically, inhibition was calculated by subtracting the average Cq value of the IAC in the no-template controls (NTCs) from the Cq value of the IAC in each well containing a wastewater or food sample. Inhibition was considered unacceptable if the Cq shift exceeded 2 cycles (37), which corresponds to 75% reduction in the target concentration if 100% PCR efficiency is assumed. The percentage of inhibition was calculated using the following formula:

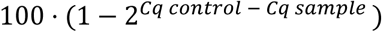

For each sample, the mean and standard deviation (SD) were calculated across subsamples, from log_10_ transformed data. Wastewater samples with an SD greater than 0.50 between subsamples were deemed invalid. For easier interpretation, standard deviations of log_10_ transformed data were also converted to geometric coefficients of variation (%CV) on the linear scale, by using the following formula (38):

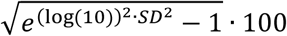

The limit of blank, i.e. the lowest concentration at which a sample was to be considered positive, was set clearly above the highest concentration obtained in the NTC and NEC wells included in the study.

In addition, for the wastewater samples, during 2024 and 2025, a control chart was established by using a positive control consisting of archived wastewater TNA extract, positive for PMMoV, to study run-to-run variability in RT-qPCR.

### Data analysis

The data curation and the statistical analyses were conducted in R version 4.3.1-4.4.1 (39). Plots were generated using the ggplot2 (40), ggmap (41) and ggthemes (42) packages.

#### Wastewater samples

For each WWTP, the daily PMMoV load was calculated by accounting for the daily wastewater flow, as follows:

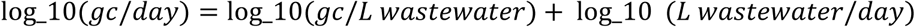

Similarly, the daily PMMoV load per resident was calculated by accounting for the number of residents connected to each WWTP (see Tables 1 and 2), as follows:

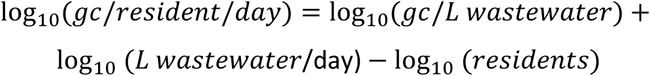

##### Short-term monitoring

To examine how PMMoV levels varied across different WWTPs (Table 1), we used a generalised additive mixed model (GAMM), hereafter referred to as Model 1, with log_10_ gc/day/resident as the response variable. WWTP was included as a fixed effect to evaluate general differences between WWTPs, while smooth terms for sampling date were fitted separately for each WWTP to capture site-specific temporal trends. To account for serial correlation in the time series data, a first-order continuous-time autoregressive correlation structure [CAR(1)] was included for each WWTP using the corCAR1 function. The model was fitted using the gamm function from the mgcv package (43) in R (31), with restricted maximum likelihood (REML) for smoothness selection. Residual autocorrelation was assessed using the acf function, and additional model diagnostics, including the k-index, residual normality and homoscedasticity, were evaluated using gam.check and diagnostic plots from mgcv. Thin plate regression splines were used for smoothing terms.

To examine the linear relationship between daily total PMMoV levels and the number of residents connected to the WWTPs, we developed a generalised least squares (GLS) model (Model 2). The response variable was the log-transformed daily PMMoV load (log_10_ gc/day), and the explanatory variable was the log-transformed number of residents (log_10_ residents) connected to the WWTPs. The model was fitted using the gls function from the nlme (44,45) package in R (31), incorporating the same autoregressive correlation structure as in the previous analysis (Model 1).

We also evaluated several alternative model variants by sequentially adding additional explanatory variables to Model 2, including the overall socioeconomic index of the municipalities covered by each WWTP catchment area, the log_10_-transformed proportion of foreign-born residents and the geographic coordinates (latitude and longitude) of the WWTPs. Geographic variables were included to assess potential spatial effects. The different models were compared using the Akaike Information Criterion (AIC), with lower values indicating a better balance between fit and model complexity. Multicollinearity among predictors was assessed using the vif function from the car (46) package in R (31).

##### Long-term monitoring

To examine how PMMoV levels varied across different WWTPs over an extended period (Table 1), we used a GAMM (Model 3) with a structure similar to Model 1 but included an additional smoothing term for the decimal day of the year to capture general seasonal trends. This seasonal component was modelled using cyclic cubic regression splines.

##### Daily monitoring

To examine how PMMoV levels varied across calendar days and between weekdays, we used a generalised additive model (GAM, Model 4) with log₁₀-transformed daily PMMoV loads (log_10_ gc/day) as the response variable. Weekday was included as a fixed effect to assess systematic differences between days of the week, while sampling date was modeled as a smooth term to capture underlying temporal trends. The model was fitted using the gam function from the mgcv (43) in R (31), with REML used for smoothness selection. Model diagnostics were performed as described above.

#### Food samples

For the food samples, PMMoV levels were converted to log_10_ gc/g (wet weight) and then adjusted to reflect PMMoV levels per typical serving size for better interpretability. These calculations were based on standard portion sizes for various food categories or, when available, serving sizes specified on food packaging, online recipes, or ingredient charts. The serving sizes for each sample are detailed in Supplementary Table 2.

## Results

### PMMoV in wastewater

#### Quality control

Untreated wastewater was collected from up to 18 WWTPs over a period of up to 34 months across three overlapping sampling series: long-term, short-term and daily monitoring (Table 1). A total of 987 samples were collected. Of these, 966 samples passed quality control and were included in the final analysis.

Inhibition monitoring started in March 2024 (Figure 2A). Overall, inhibition levels were relatively low, with an average of 0.36 Cq units, corresponding to a 29% reduction in target concentration if 100% PCR efficiency is assumed. Fifteen samples displayed inhibition exceeding 2 Cq units. These were deemed invalid and excluded from the study.

**Figure 2.**
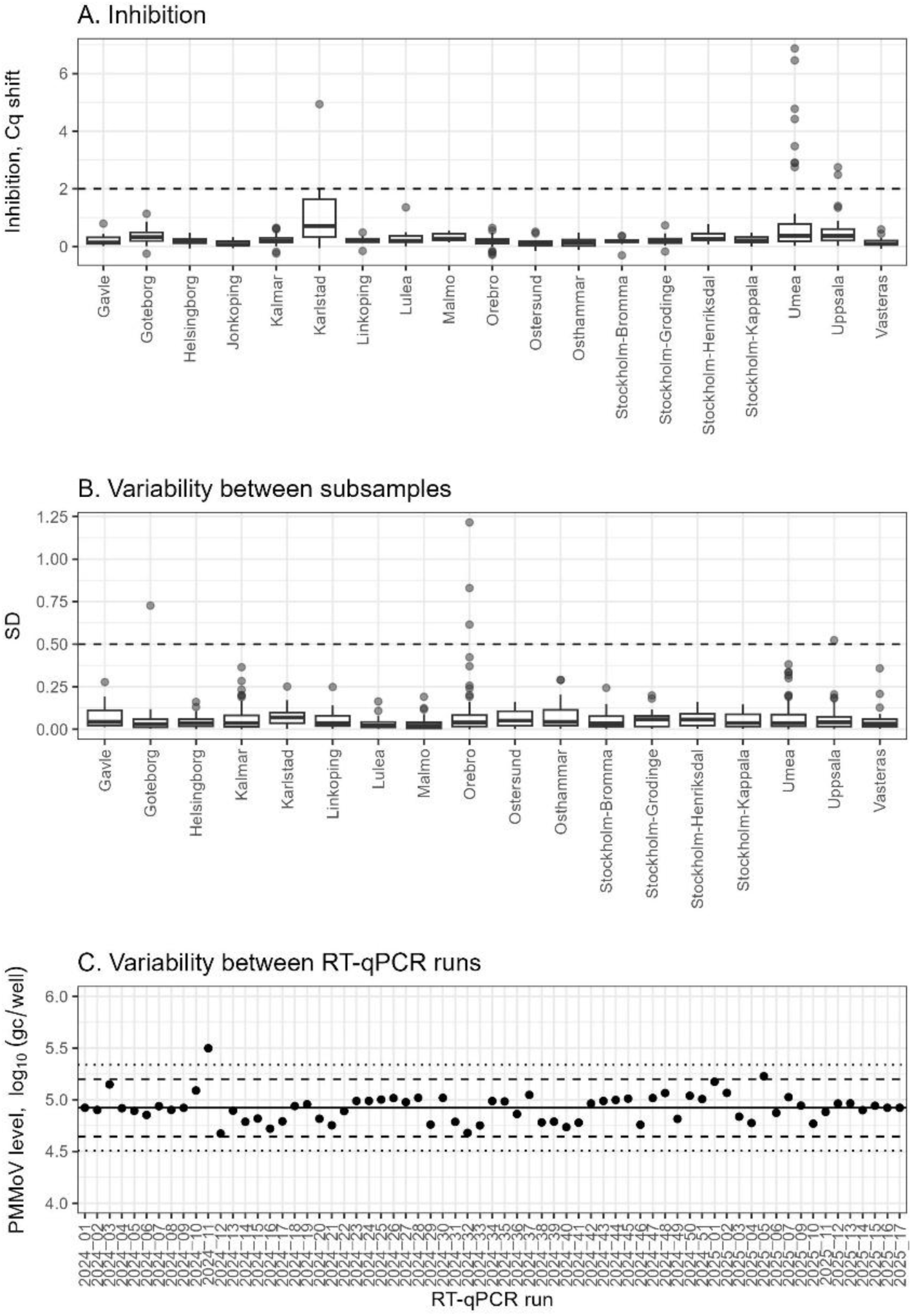
Quality control data from wastewater analyses. **A.** RT-qPCR inhibition for the wastewater samples at each WWTP, with the dotted line indicating the threshold for acceptable inhibition. **B**: SD between subsamples (for log_10_ transformed data), with the dotted line indicating the threshold for acceptable SD. **C:** Control chart for RT-qPCR during 2024 and 2025, based on a positive control of RNA extracts of PMMoV positive wastewater. The black dots represent the value of the positive controls, the solid black line represents the overall mean of these controls and the dotted lines represents the mean +/-2・SD and +/-3・SD, respectively

Each sample was extracted in two subsamples. Between subsamples, for log_10_ transformed data, the average SD was 0.061, corresponding to a geometric CV of 14% on the linear scale. Five samples exhibited an SD greater than 0.50 and were consequently deemed as invalid (Figure 2B).

Out of 622 negative control wells (NTCs and NECs) used for the three wastewater monitoring series, 189 showed a positive signal in RT-qPCR. While most signals were detected at very low concentrations, the highest value was 9 copies per reaction for the NTCs and 120 copies per reaction for the NECs. The limit of blank was set at 150 copies per reaction. One sample (Kalmar week 25, 2023) had a concentration below the limit of blank and was deemed invalid.

A control chart was established on week 1 in 2024 to assess variability between different RT-qPCR runs during the short-term and long-term monitoring (Figure 2C). Between runs, the measured concentration of the positive control fluctuated between 4.67 and 5.50 log_10_ gc/well. The between-run SD was 0.14, corresponding to a geometric CV of 33% on the linear scale.

Supplementary Figures 1 and 2 provide flow data (L/resident/day) and measured PMMoV levels (gc/L and gc/resident/day) for all valid samples throughout the study.

#### Spatial variation in PMMoV levels and relation between total PMMoV levels and number of residents (short-term monitoring)

Over six months of weekly monitoring, PMMoV levels remained generally stable both within and across the 18 WWTPs included in the study (Model 1, Figure 3A-B). The model’s adjusted *R²* was relatively low (0.23), indicating that only a small portion of PMMoV variability was explained by city-specific differences or temporal trends. The average PMMoV concentration across all sites was 10.37 log_10_ gc/inhabitant/day, with Karlstad exhibiting the highest predicted mean and Östhammar the lowest; a difference of 0.35 log_10_ units. For comparison, PMMoV concentrations expressed in gc/L are shown in Figure 3C, with an overall mean of 7.86 log_10_ gc/L.

**Figure 3.**
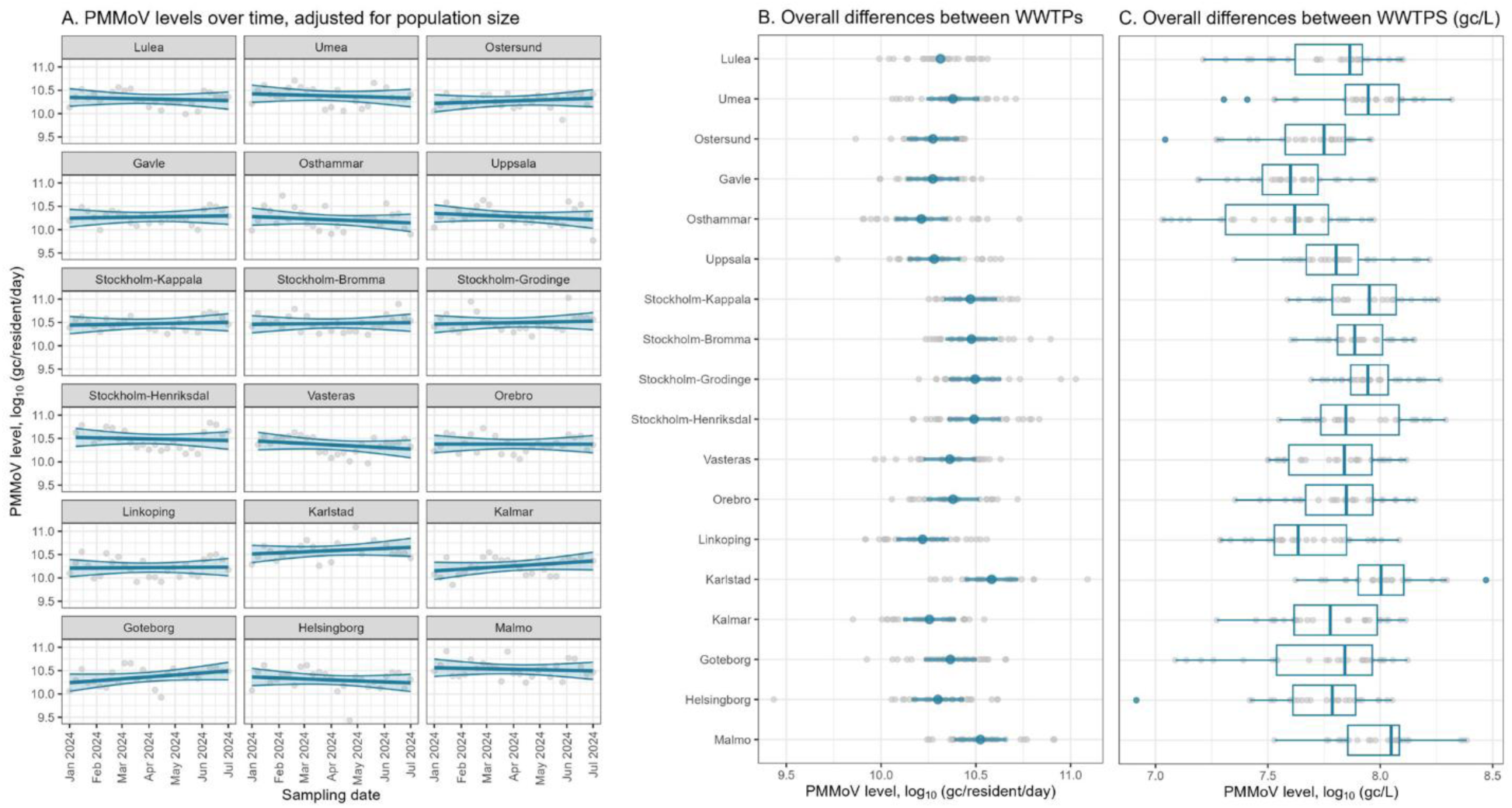
Results from the short-term monitoring in 18 WWTPs. **A**. Time series of measured values for each WWTP, with fitted GAMM smooths (Model 1) showing temporal trends during the sampling period, along with their 95% confidence bands. Grey dots represent individual data points. **B**. Effect of each WWTP from Model 1. The blue points represent the estimated effects, while the horizontal bars indicate 95% confidence bands. Luleå does not have a confidence band, as it was used as the reference category in the model, meaning the effects of other sites are interpreted relative to it. The WWTPs are plotted in a north to south direction. Grey dots represent individual data points. **C**. Measured PMMoV concentrations, expressed in log_10_ gc/L, for comparison

A linear relationship was observed between the total log_10_ PMMoV load and the log_10_ population served by each WWTP, though with high residual variability (Model 2, Figure 4). To explore other potential influences on PMMoV excretion, we also included data on population structure, specifically the percentage of foreign-born residents and a socioeconomic index for each municipality served by the WWTPs (Figure 5), as these factors may affect dietary habits. Additionally, latitude and longitude of the WWTPs were also collected, based on geographic trends in PMMoV-levels reported by Rosengart *et al.* (10). Each variable was assessed separately to Model 2, but none improved model fit and were therefore excluded from the final version. This suggests that population structure and geographic differences are not major contributors to PMMoV variation in wastewater across the WWTPs included in this study. Detailed parameter estimates, *p*-values and AIC values for all tested models are provided in Supplementary Table 3.

**Figure 4.**
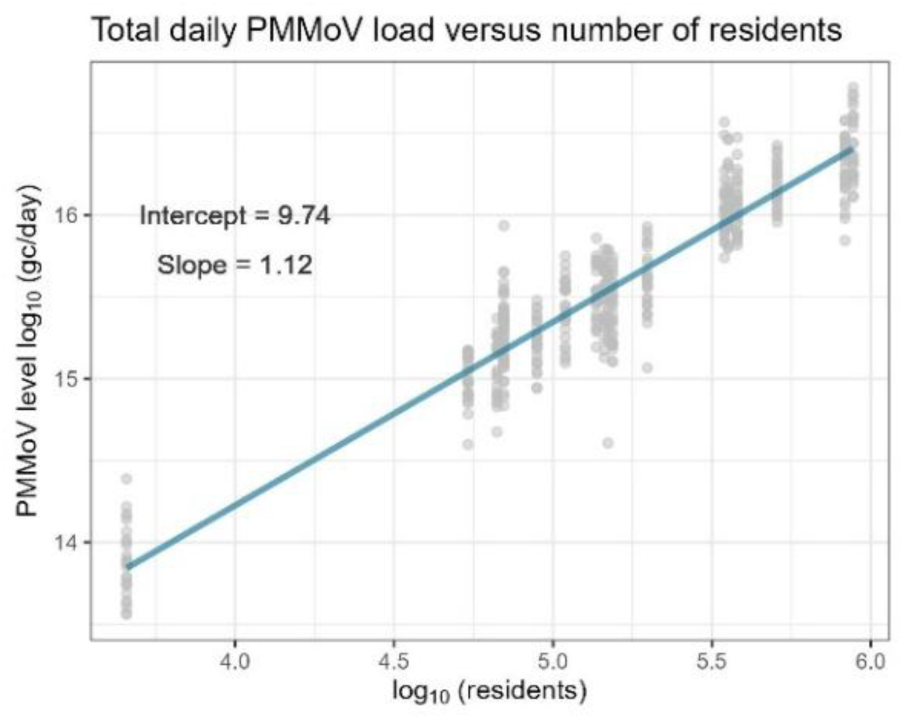
Linear relationship between total daily PMMoV levels and the number of residents connected to the WWTPs. Grey dots represent individual data points, while the solid line indicate the predicted means from a GLS regression model (Model 2)

**Figure 5.**
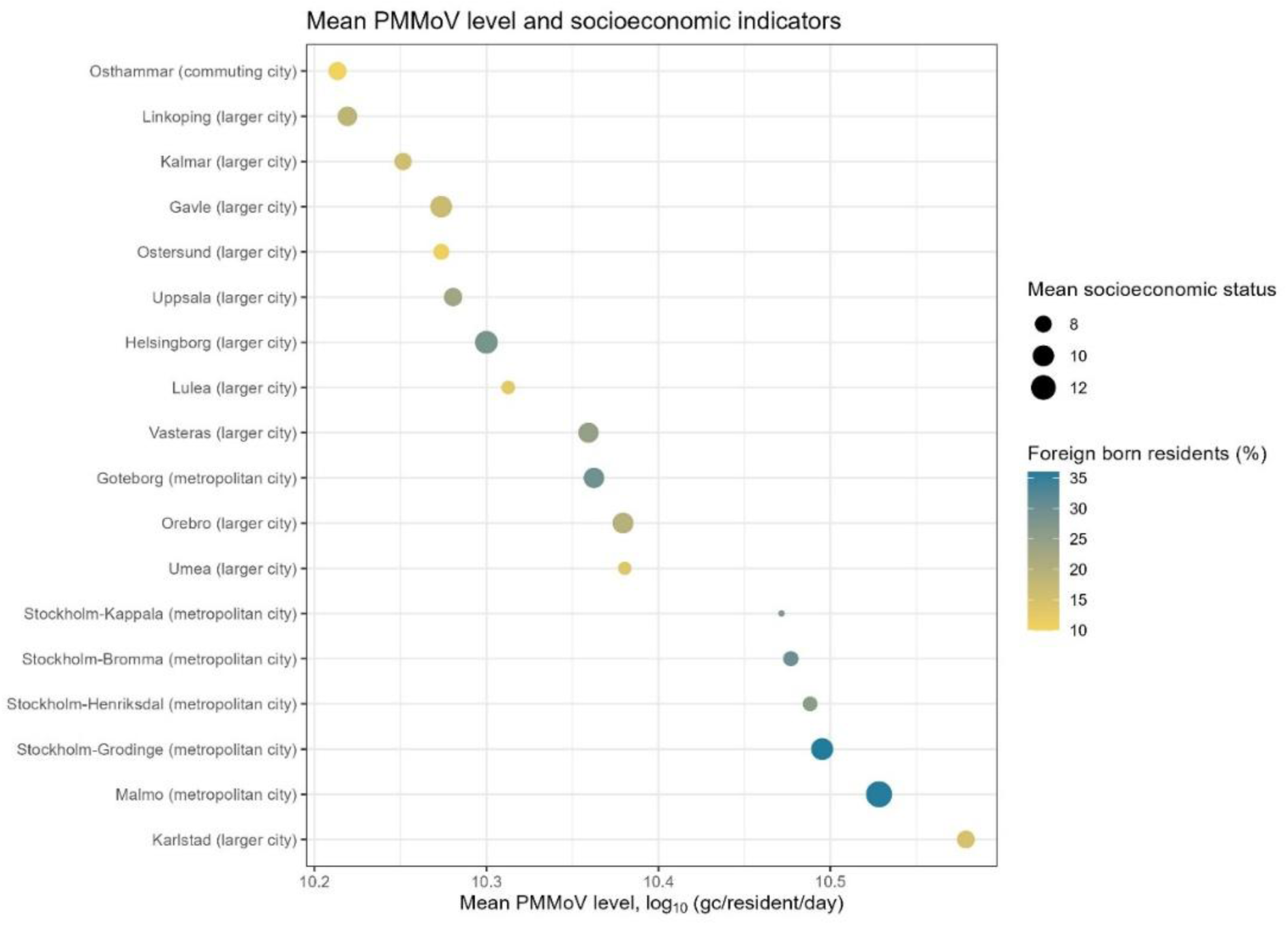
Mean per-resident PMMoV levels from each WWTP, arranged from lowest to highest, alongside socioeconomic status and the percentage of foreign-born residents within the municipalities covered by the WWTP catchment area. The socioeconomic index combines the percentage of inhabitants with pre-secondary education or less, low economic standards and reliance on economic assistance or long-term unemployment. Higher values (larger dots) indicate poorer socioeconomic status. The official classification of each city is indicated in parenthesis on the y-axis

#### Long-term temporal variation in PMMoV levels (long-term monitoring)

During 34 months of weekly sampling across four geographically distinct WWTPs, PMMoV levels remained generally stable over time, though some fluctuations were observed, especially for Kalmar, but no consistent temporal trends were identified (Figure 6A). Predicted mean PMMoV levels ranged from 9.97 to 10.37 log_10_ gc/resident/day. To investigate potential seasonal trends, we incorporated a seasonal component into the model, but no pronounced overall seasonal pattern was detected (Figure 6B). The adjusted *R²* of the model was 0.19.

**Figure 6.**
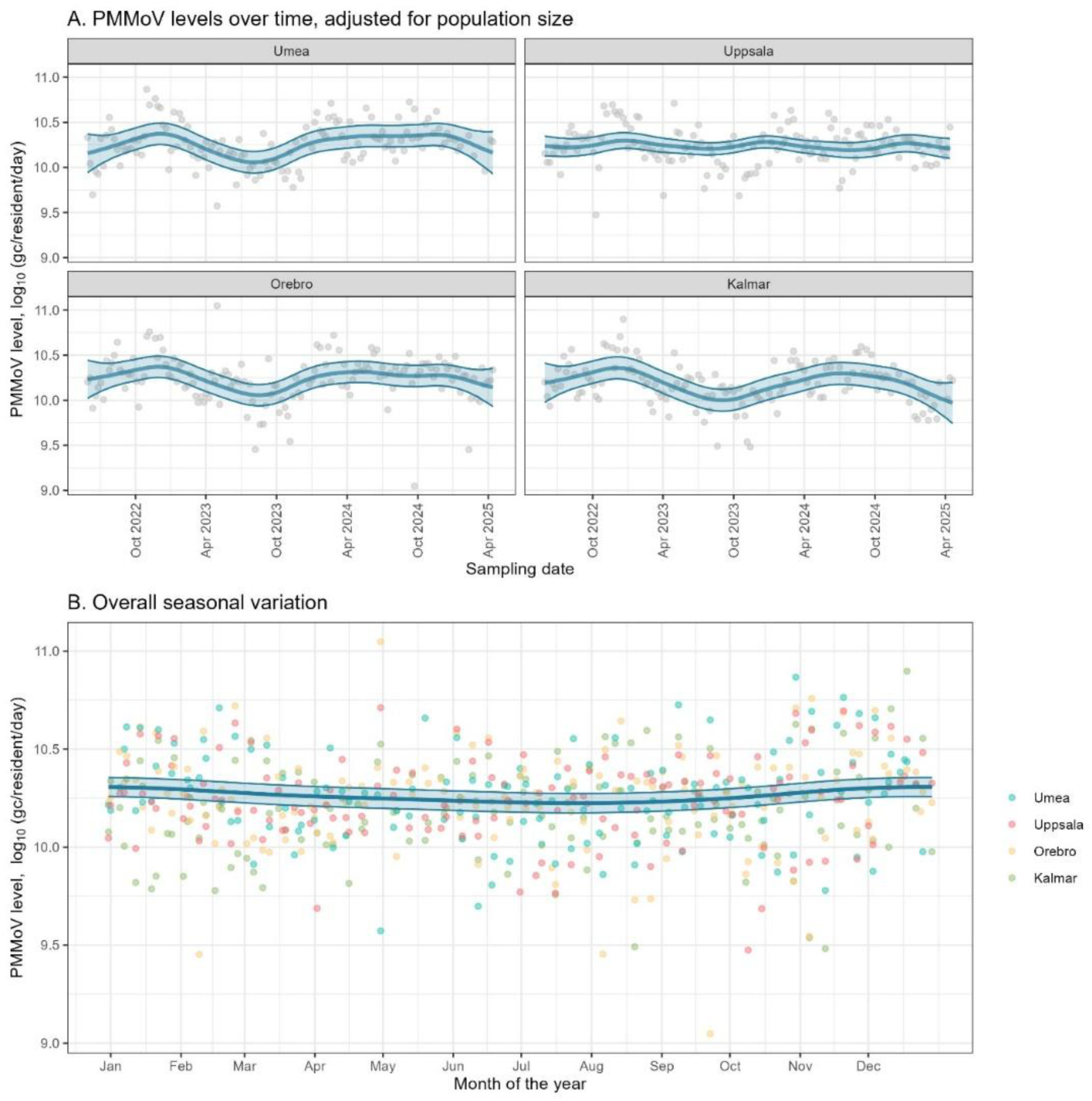
Results from the long-term monitoring in four WWTPs. **A.** Trends in PMMoV levels over time for each WWTP, with dots representing individual data points, solid lines indicating predicted means and shaded areas showing 95% confidence bands from GAMM smooths (Model 2). **B**. Overall seasonal variation in PMMoV levels across the sampling period, with individual observations for each WWTP (dots) and a predicted mean with a 95% confidence band from Model 2

We also examined the relationship between PMMoV levels (log_10_ gc/resident/day) and wastewater temperature (°C) using a GLS regression model with the data from 2024. In that year, daily average temperatures ranged from 8 to 21°C (Supplementary Figure 3). However, no significant linear association was found (intercept = 10.35; slope = -0.00025; *p*-value associated with the slope estimate = 0.64), indicating that temperature variations within this range are unlikely to have a meaningful impact on PMMoV levels.

#### Day-to-day variation in PMMoV levels (daily monitoring)

Daily sampling of PMMoV over five weeks at a WWTP in Uppsala showed relatively stable PMMoV levels across days, with no pronounced association between PMMoV levels and specific weekdays (Model 4, Figure 7). The adjusted *R^2^* of the model was 0.68.

**Figure 7.**
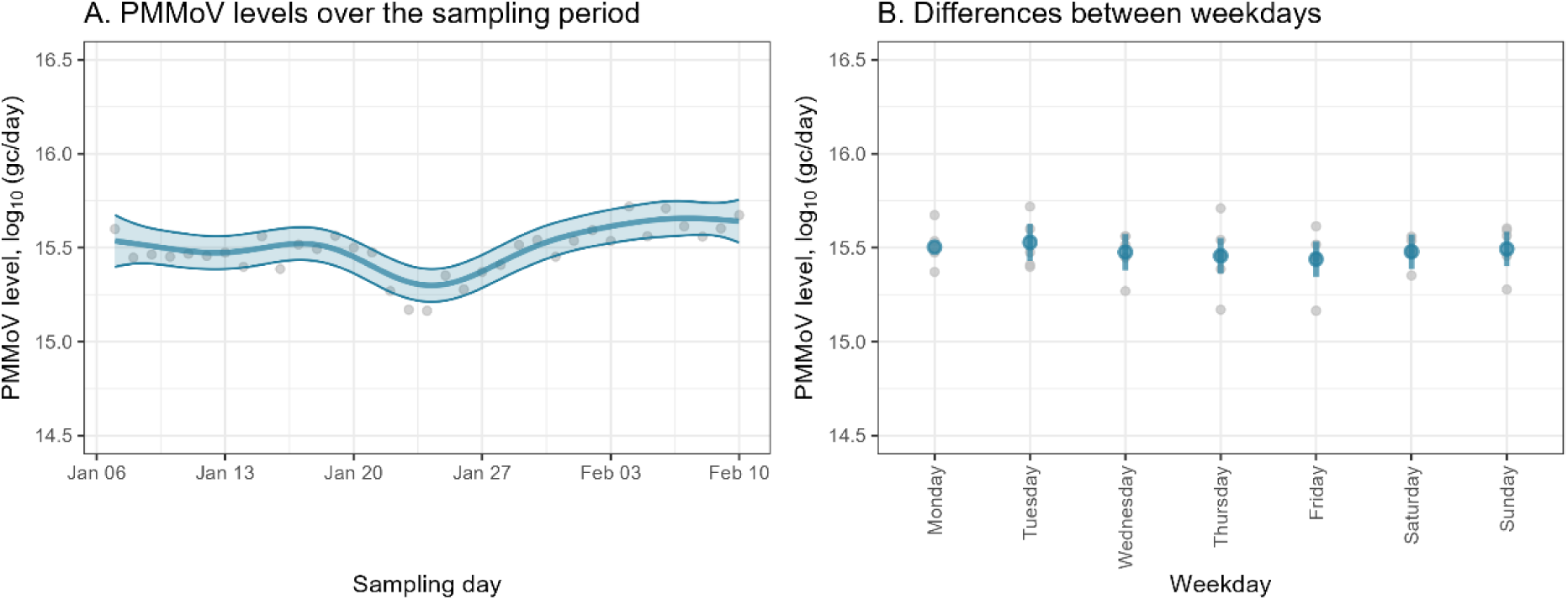
A. Time series of measured PMMoV levels each day, with a fitted GAM smooth (Model 4) showing trends during the sampling period, along with their 95% confidence bands. Dots represent individual data points. **B.** PMMoV levels at different weekdays. The blue dots represent the estimated effects, while the bars indicate 95% confidence bands. Monday does not have a confidence band, as it was used as the reference category, meaning the effects of other days are interpreted relative to it. Grey dots represent individual data points

### PMMoV in food products

We analysed various food items to identify potential dietary sources of PMMoV. Of the 60 food products tested, 45 exhibited detectable levels of PMMoV, surpassing the limit of blank (set at 150 copies per reaction, consistent with the threshold established in the wastewater study). None of the negative controls (NTCs or NECs) used in the food analyses exceeded this limit, and no food sample showed more than 2 Cq values of inhibition.

High PMMoV levels were detected across a range of food products, particularly in spices, spice mixes, premade and processed foods, pre-made sauces and various snack-related items. The highest concentrations were found in pre-made vegetarian chicken, a crisp dip seasoning blend and paprika powder, ranging from 12.21 to 11.31 log_10_ gc/serving (Figure 8). Notably, two of the three *Capsicum* fruits (paprika and spitzpaprika) had levels below the limit of blank, yet all tested spices and spice mixes, and the majority of the pre-made foods, both those sold fresh and frozen, exhibited high PMMoV levels. The estimated serving sizes and PMMoV levels expressed in copies per gram are provided in Supplementary Table 2.

**Figure 8.**
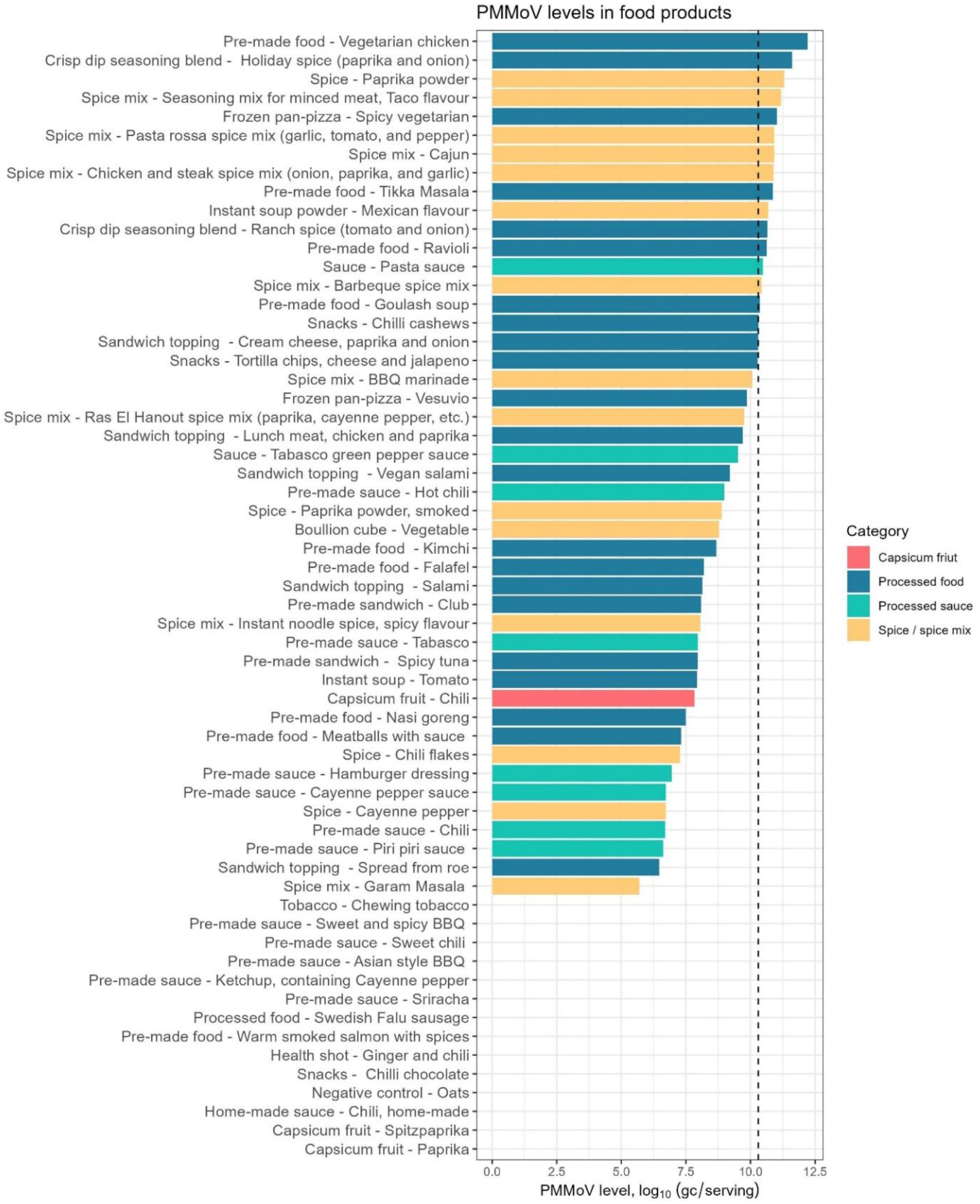
PMMoV levels in various food products, presented as gc/serving. As a reference, the dotted vertical line indicates the average daily per-resident PMMoV level in wastewater (10.37 log_10_ gc/resident/day), as determined from the short-term wastewater monitoring studys

Assuming that PMMoV remain largely intact as it passes through the digestive tract without significant degradation, and, conversely, that there is no substantial breakdown of plant cell walls or viral particles that could make the RNA more easily detectable in faeces than in food, our results suggest that a single serving or even significantly less of several tested foods could potentially account for the average daily PMMoV levels detected in faeces (10.37 log_10_ gc/resident/day). However, a potential discrepancy between PMMoV levels in food and wastewater may stem from differences in extraction efficiencies between the methods used for their analysis. To assess this, we conducted a control extraction on three food items using the wastewater extraction method. The results closely matched those obtained with the plant extraction method (Supplementary Table 4).

## Discussion

This study aimed to evaluate the presence and variability of PMMoV in raw wastewater across different locations and over time in Swedish WWTPs, as well as to explore its potential dietary sources to better understand its origin in domestic wastewater. Together, these insights contribute to a better assessment of the suitability of PMMoV as a human faecal marker in wastewater.

First, with regards to analytical method performance in the wastewater study, we observed generally low inhibition and low variation between subsamples (replicate extractions) (Figure 1A-B). However, greater variation was evident between separate RT-qPCR runs. Since the introduction of a control chart (Figure 1C), we observed a weak positive correlation between the positive control value and the average estimated PMMoV concentration in wastewater samples (log₁₀ gc/resident/day) across WWTPs on each plate (Pearson’s *r* = 0.39, *p* = 0.0013). This suggests that technical variability between runs may partially account for the temporal fluctuations in PMMoV levels observed over time. Transitioning from qPCR to digital PCR (dPCR) may reduce this effect, as dPCR typically provides more consistent quantification across runs (34). For example, a previous study on hepatitis A virus demonstrated a reduction in geometric CV from 35% between RT-qPCR runs (comparable to the 34% variation observed in this study) to 1.2% with RT-dPCR (35).

During the short-term monitoring of 18 WWTPs across Sweden, PMMoV concentrations were generally stable, both temporally within individual WWTPs and spatially between plants (Figure 3). No clear associations were observed between PMMoV levels and socioeconomic indicators, population composition (proportion of foreign-born residents), or geographical location (Figure 5). These results are consistent with an earlier study in Kentucky, USA, which found no effect of household income on PMMoV concentrations in wastewater (21). Together, these results support the suitability of PMMoV as a broadly applicable marker for human faeces in WBS.

Among the WWTPs, the largest difference in mean PMMoV concentration was observed between Östhammar and Karlstad, a 0.35 log_10_ difference, which corresponds to a 2.34-fold difference on a linear scale. This difference may reflect differences in the wastewater matrix unique to each plant. For example, while Karlstad had similar flow rates to other WWTPs, it exhibited both higher PMMoV concentrations (gc/L; Figure 3C) and slightly greater inhibition (Figure 1), potentially suggesting a distinct matrix composition. Such differences highlight the importance of PMMoV not only as a population marker but also as a process control in WBS.

We also investigated whether certain industrial discharges could influence PMMoV values. For example, one of Sweden’s largest crisp manufacturers, which produces paprika and chilli products, is connected to the WWTP in Göteborg. However, the PMMoV concentrations in Göteborg did not stand out from those of the other WWTPs, suggesting that any contribution from the food industry was likely diluted due to the large population it supplies. No similarly large food industries are known to be connected to the Karlstad treatment plant.

In general, the PMMoV levels we detected, both in gc/L (on average 7.86 log₁₀ gc/L) and gc/resident/day (on average 10.37 log₁₀ gc/resident/day), are consistent with the results of other studies, including those in the United States [(21); approximately 6–10 log_10_ gc/L], Canada [(22); ∼9–11 log_10_ gc/resident/day and (11); ∼7–8 log_10_ gc/L], Germany [(23); ∼5–7 log_10_ gc/L] and Saudi Arabia [(24); ∼5–7 log_10_ gc/L], based on data from figures in the corresponding articles).

In terms of geographic location, a large study from the United States reported longitudinal variation in PMMoV concentrations, with higher levels in western regions than in the east (10). While our study covered a smaller geographical area, no such regional trend was observed within Sweden. Despite spanning four climatic zones, we found no evidence that latitude, temperature, or seasonal differences influenced PMMoV concentrations.

As expected, we observed a strong linear relationship between the log_10_ daily PMMoV load and the log_10_ number of residents connected to each WWTP across a wide range of population sizes (Figure 4). This further supports the use of PMMoV as a proxy for population size in WBS. A similar relationship has been reported in a study from Missouri, USA, which covered a comparable range of WWTP catchment sizes (47).

Next, during long-term monitoring of four geographically separated WWTPs over nearly three years (Figure 6), we observed no consistent seasonal patterns in PMMoV levels, nor any clear association with wastewater temperature. This apparent stability aligns with a previous experimental study demonstrating the resilience of PMMoV across a wide temperature range (48). To date, only a limited number of long-term studies (spanning two years or more) have been published (11,22).

Temporal variations in WBS data may reflect changes in the number of people contributing to the system, which is one of the main motivations for using PMMoV in WBS, in addition to its role as an analytical control. Mobile data on population mobility in Sweden shows that population numbers in larger cities such as Kalmar, Örebro, Uppsala and Umeå are relatively stable throughout the year, except in July (the main holiday season) and around Christmas (49). In July, these cities usually record a population decline of around 10%. However, this seasonal dip was not clearly visible in the PMMoV data (Figure 6; Supplementary Figure 2). Future studies could improve temporal resolution by increasing sampling frequency, refining quantification methods (e.g. with dPCR) and comparing PMMoV trends directly with mobility data, such as mobile phone mobility data (50). Such approaches would further validate PMMoV as a marker for population size in WBS.

Since PMMoV is ingested through food and certain foods containing ingredients derived from *Capsicum* species, such as chilli-flavoured snacks and tacos, tend to be consumed more frequently on certain days, we hypothesised that there might be day-to-day differences in PMMoV concentrations in wastewater, which, if present, would affect the reliability of PMMoV as a faecal marker. However, our daily monitoring showed no clear differences in PMMoV concentrations on the different days of the week (Figure 7). A recent study from the United States also reported low daily variability in PMMoV concentrations in raw wastewater, with fluctuations comparable to other biological markers less strongly associated with diet (the cryptic plasmid pBI143 and CrAssphage) (51).

A central aim of this study was to identify dietary sources of PMMoV. For PMMoV to be a reliable human faecal indicator, it must be consistently present in commonly consumed foods. Our findings show that even a single serving, or much less, of several tested foods could theoretically account for the average daily PMMoV levels observed in wastewater (Figure 8). Paprika powder, for example, contained some of the highest concentrations (11.25 log_10_ gc/g), suggesting that as little as 0.1 g could match the average daily faecal shedding per resident. Given its widespread use as a flavouring and colouring agent in processed foods, this may partly explain the persistently high PMMoV levels in human wastewater.

Detected levels were in line with, or exceeded, those reported in previous studies, which have mostly focused on spicy ingredients. While earlier work has documented levels of up to 8 log_10_ gc/g or mL (15,26,27), few studies have assessed processed, milder or ready-made foods. One exception is Zheng et al. (15) who analysed full meals, though without specifying the foods tested.

Importantly, PMMoV-infected *Capsicum* plants are more likely to be processed into sauces or spices than sold fresh, as the infection often affects the appearance of the fruit (52). PMMoV is also known to withstand standard food processing methods (15). Consequently, spices and processed foods are likely to contribute more significantly to PMMoV levels in human faeces than fresh *Capsicum* fruits. Our results confirm this, as the highest PMMoV concentrations were found in spices and various processed foods, while only one of three *Capsicum* fruits had detectable PMMoV levels.

Our food analysis has several limitations. Serving sizes vary between individuals, introducing uncertainty to the values shown in Figure 8, although copies per gram are provided in Supplementary Table 2. We also tested only a single sample per food type, and PMMoV levels are likely to vary between batches of the same product. Furthermore, differences in extraction efficiency across food matrices cannot be fully excluded. Despite these limitation, this study remains the most comprehensive investigation to date of dietary PMMoV sources and provides valuable insight into the foods contributing to its high prevalence in human faeces.

## Conclusions

We found that PMMoV is consistently present at high levels in raw wastewater across WWTPs of varying sizes, population compositions and geographic locations in Sweden. A linear relationship between total daily PMMoV load and the number of residents served by each WWTP underscores the potential of PMMoV as a population marker in WBS.

Our results also demonstrate that PMMoV levels in wastewater remain relatively stable over time, showing no major seasonal fluctuations or temperature-related effects.

Exploratory analysis of food products revealed a notable presence of PMMoV in commonly consumed items, including spices and a range of processed foods such as sauces, sandwich spreads, snacks and ready-made meals. These dietary sources likely explain the consistent detection of PMMoV in human wastewater across diverse locations and seasons.

In summary, this study provides a comprehensive characterisation of PMMoV as a faecal indicator and population normaliser, while providing valuable insights into its dietary origins. Our findings regarding PMMoV in wastewater are in agreement with those reported in similar international studies.

## Author contributions

Conceptualisation: SP, AJS; Methodology: SP, NM; Software: SP; Validation: SP; Formal analysis: SP; Investigations: SP, JEV, SC, FPer, FPet, LD, PV, MV, NM, AJS; Recourses: AJS; Data curation: SP, JEV, SC; Writing - Original Draft: SP, AJS; Visualisation: SP; Supervision: AJS; Project administration: AJS, MM, Writing, Review, and Editing: All authors.

## Data Availability

All data produced in the present study are available upon reasonable request to the authors

## Acknowledgement

We sincerely thank all participating WWTPs for their invaluable efforts in collecting samples and associated data throughout the study. We are especially grateful to Susanne Tumlin of Gryaab AB for her suggestions and constructive feedback on the manuscript. Our thanks also go to Katalin Nemes at the Swedish Food Agency for her advice on food and plant extraction methodologies, and to Claudia von Brömssen from SLU for her statistical advice. This study was funded by Carl Trygger’s Foundation for Scientific Research (CTS 22:2255), the SciLifeLab Pandemic Laboratory Preparedness Programme (grants C19RA:031, LPP1:008 and REPLP1:007) and governmental grants provided to the Public Health Agency of Sweden under assignment S2024/00187.

