## Supplementary Material for "Assessing PMMoV as a faecal marker for wastewater-based surveillance - Insights from Swedish wastewaters and foods"

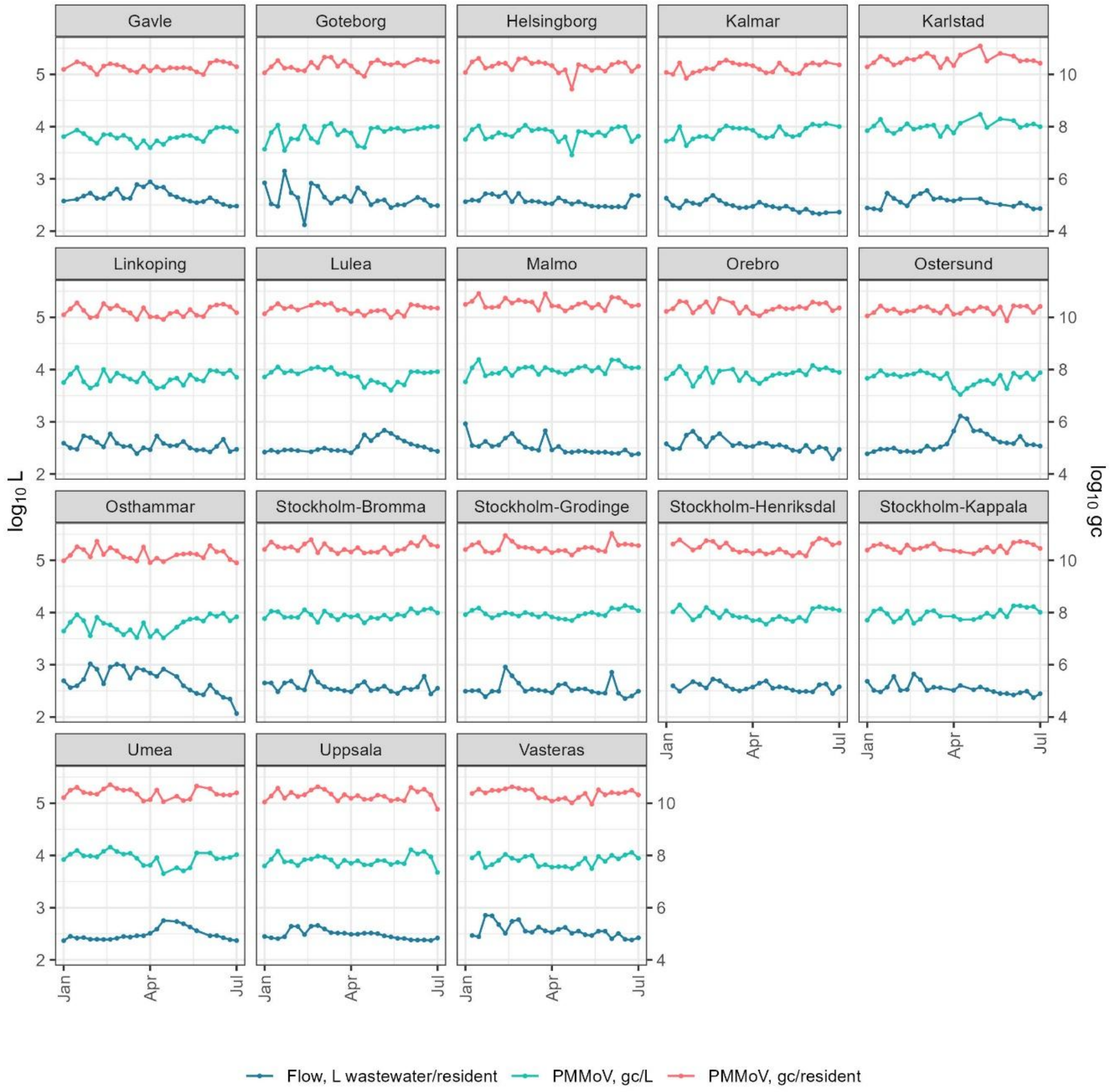


***Supplementary Figure 1****. Valid measurements from the short-term monitoring of raw wastewater, showing daily wastewater flow per resident and PMMoV content (gc/L and gc/resident/day)*

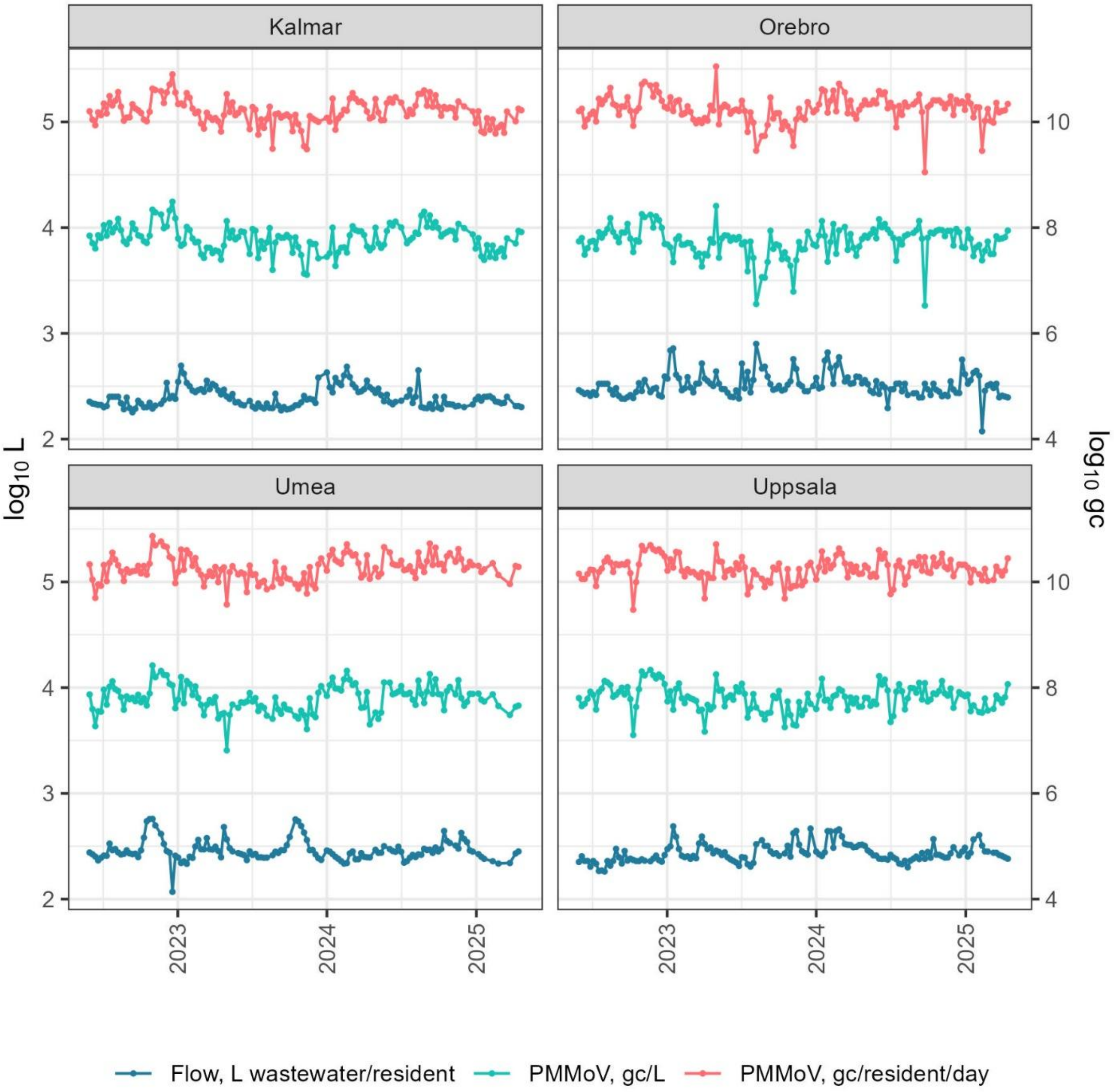


***Supplementary Figure 2.*** *Valid measurements from the long-term monitoring of raw wastewater, showing daily wastewater flow per resident and PMMoV content (gc/L and gc/resident/day)*

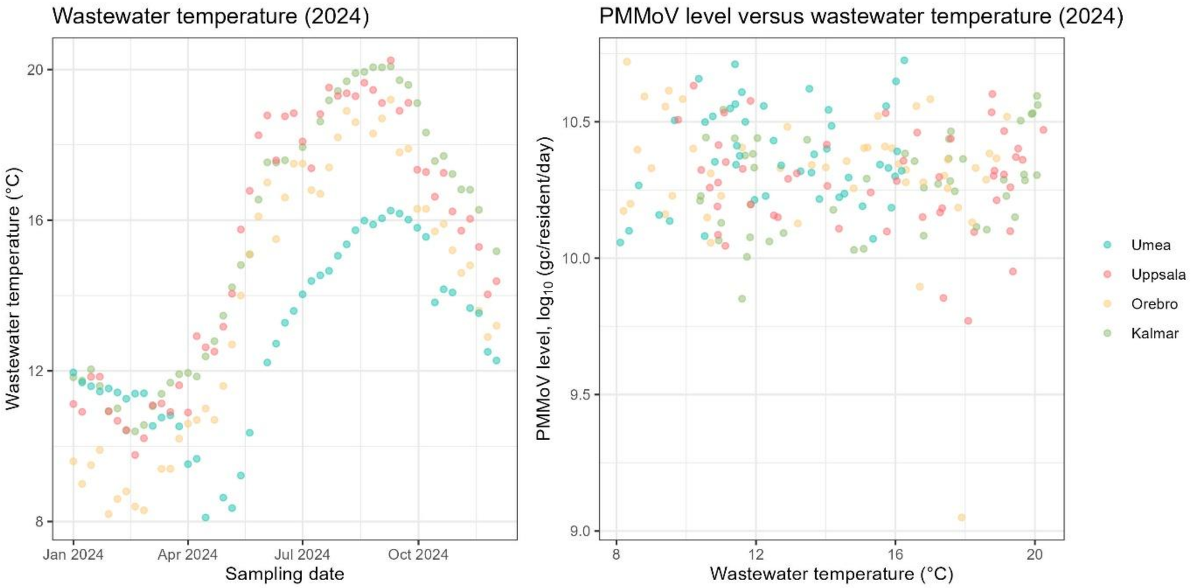


***Supplementary Figure 3**. Left: daily average wastewater temperature for 2024 for the WWTPs involved in the long-term monitoring. Right: PMMoV level versus daily average wastewater temperature*

***Supplementary Table 1.*** *List of samples with missing flow values*

| Sampling date | WWTP |
|---|---|
| 07-18-2022 | Kalmar |
| 07-18-2022 | Örebro |
| 07-25-2022 | Kalmar |
| 07-25-2022 | Örebro |
| 08-01-2022 | Kalmar |
| 08-01-2022 | Örebro |
| 08-08-2022 | Kalmar |
| 08-08-2022 | Örebro |
| 01-01-2024 | Helsingborg |
| 01-01-2024 | Stockholm-Grödinge |
| 01-08-2024 | Stockholm-Käppala |
| 01-15-2024 | Malmö |
| 01-29-2024 | Malmö |
| 01-29-2024 | Stockholm-Grödinge |
| 02-05-2024 | Gävle |
| 02-05-2024 | Stockholm-Grödinge |
| 02-05-2024 | Stockholm-Käppala |
| 02-12-2024 | Örebro |
| 02-19-2024 | Helsingborg |
| 03-04-2024 | Gävle |
| 03-04-2024 | Helsingborg |

| | |
|---|---|
| 03-04-2024 | Stockholm-Grödinge |
| 03-04-2024 | Stockholm-Käppala |
| 03-04-2024 | Västerås |
| 03-11-2024 | Gävle |
| 03-18-2024 | Helsingborg |
| 03-25-2024 | Örebro |
| 04-29-2024 | Helsingborg |
| 05-20-2024 | Malmö |
| 05-20-2024 | Västerås |
| 05-27-2024 | Stockholm-Bromma |
| 05-27-2024 | Stockholm-Henriksdal |
| 05-27-2024 | Västerås |
| 06-17-2024 | Linköping |
| 06-17-2024 | Östersund |
| 06-24-2024 | Östersund |
| 07-01-2024 | Stockholm-Grödinge |
| 07-15-2024 | Kalmar |
| 07-15-2024 | Örebro |
| 08-05-2024 | Kalmar |
| 08-05-2024 | Örebro |
| 09-23-2024 | Kalmar |
| 09-23-2024 | Örebro |
| 10-14-2024 | Kalmar |

| | |
|---|---|
| 10-14-2024 | Örebro |
| 01-13-2025 | Kalmar |
| 01-13-2025 | Örebro |
| 01-20-2025 | Uppsala |
| 02-03-2025 | Kalmar |
| 03-03-2025 | Umeå |

*Supplementary Table 2. Details of tested food items.*

| Sample | Name | Description | Category | Brand/origin | Part of food | Proccessing method | Estimated typical serving size (g) | PMMoV concentration ($log_{10}$ copies/g) |
|---|---|---|---|---|---|---|---|---|
| 1 | Chilli chocolate | Snacks | Processed food | Lindt | Homogenised | Grinding with a mortar and pestle | < LOB. not estimated | < LOB |
| 2 | Mexican flavour | Instant soup powder | Spice / spice mix | Knorr | Homogenised | Grinding with a mortar and pestle | 16 | 9.48 |
| 3 | Spread from roe | Sandwich topping | Processed food | Ejdern | Homogenised | Vortex mixing in buffer | 12 | 5.39 |
| 4 | Holiday spice (paprika and onion) | Crisp dip seasoning blend | Processed food | Estrella | Homogenised | Vortex mixing in buffer | 6.5 | 10.80 |
| 5 | Ranch spice (tomato and onion) | Crisp dip seasoning blend | Processed food | Estrella | Homogenised | Vortex mixing in buffer | 6.5 | 9.84 |
| 6 | Cream cheese, paprika and onion | Sandwich topping | Processed food | Tine | Homogenised | Vortex mixing in buffer | 20 | 8.99 |
| 7 | Ginger and chili | Health shot | Processed food | Rå | Homogenised | Vortex mixing in buffer | < LOB. not estimated | < LOB |
| 8 | Tomato | Instant soup | Processed food | Kelda | Homogenised | Vortex mixing in buffer | 200 | 5.63 |
| 9 | Spicy vegetarian | Frozen pan-pizza | Processed food | Garant | Homogenised toppings | Fine chopping with sterile scalpels | 50 | 9.33 |
| 10 | Vesuvio | Frozen pan-pizza | Processed food | Garant | Homogenised toppings | Fine chopping with sterile scalpels | 50 | 8.15 |
| 11 | Spicy tuna | Pre-made sandwich | Processed food | Fresh Food | Homogenised toppings | Fine chopping with sterile scalpels | 50 | 6.25 |
| 12 | Club | Pre-made sandwich | Processed food | Fresh food | Homogenised toppings | Fine chopping with sterile scalpels | 50 | 6.38 |
| 13 | Vegetarian chicken | Pre-made food | Processed food | Hot food stand, No chick Sherlock | Homogenised | Fine chopping with sterile scalpels | 125 | 10.11 |
| 14 | Warm smoked salmon with spices | Pre-made food | Processed food | Hot food stand, Smokin Salmon | Homogenised | Fine chopping with sterile scalpels | < LOB. not estimated | < LOB |
| 15 | Falafel | Pre-made food | Processed food | Hot food stand, Frikkin' falafel | Homogenised | Fine chopping with sterile scalpels | 125 | 6.10 |
| 16 | Nasi goreng | Pre-made food | Processed food | Hot food stand, Nasi goreng | Homogenised | Fine chopping with sterile scalpels | 125 | 5.40 |
| 17 | Meatballs with sauce | Pre-made food | Processed food | Hot food stand, Meatballs | Homogenised | Fine chopping with sterile scalpels | 125 | 5.22 |
| 18 | Tikka Masala | Pre-made food | Processed food | Hot food stand, Tikka Masala | Homogenised | Fine chopping with sterile scalpels | 125 | 8.77 |
| 19 | Ravioli | Pre-made food | Processed food | Tradizionali Rana | Homogenised filling | Fine chopping with sterile scalpels | 200 | 8.32 |

| | | | | | | | | |
|---|---|---|---|---|---|---|---|---|
| 20 | Vegan salami | Sandwich topping | Processe d food | Eldorado, Veganks Salami | Homogenise d | Fine chopping with sterile scalpels | 10 | 8.20 |
| 21 | Salami | Sandwich topping | Processe d food | Deliskivor, Salami Chipotle | Homogenise d | Fine chopping with sterile scalpels | 10 | 7.13 |
| 22 | Lunch meat, chicken and paprika | Sandwich topping | Processe d food | Aladin, Mortadella Kyckling paprika Skivad | Homogenise d | Fine chopping with sterile scalpels | 10 | 8.70 |
| 23 | Goulash soup | Pre-made food | Processe d food | Garant, Gulaschsopp a | Homogenise d | Vortex mixing in buffer | 200 | 8.06 |
| 24 | Kimchi | Pre-made food | Processe d food | Delief, Kimchi | Homogenise d | Fine chopping with sterile scalpels | 40 | 7.08 |
| 25 | Chilli cashews | Snacks | Processe d food | Pick&Mix, Chili Cashew | Homogenise d | Grinding with a mortar and pestle | 50 | 8.61 |
| 26 | Tortilla chips, cheese and jalapeno | Snacks | Processe d food | Santa Maria, tortilla chips, cheese &jalapeño | Homogenise d | Grinding with a mortar and pestle | 46.25 | 8.60 |
| 27 | Paprika | Capsicum fruit | Capsicu m friut | Nature choice, Röd Palermo paprika, Spain | Homogenise d | Liquid nitrogen and grinding with a mortar and pestle | < LOB. not estimate d | < LOB |
| 28 | Seasoning mix for minced meat, Taco flavour | Spice mix | Spice / spice mix | Santa Maria, Taco original | Homogenise d | Vortex mixing in buffer | 7 | 10.34 |
| 29 | Chewing tobacco | Tobacco | Spice / spice mix | VELO, Portion Snus | Homogenise d | Fine chopping with sterile scalpels | < LOB. not estimate d | < LOB |
| 30 | Paprika powder | Spice | Spice / spice mix | Santa Maria | Homogenise d | Vortex mixing in buffer | 1.13 | 11.25 |
| 31 | Cajun | Spice mix | Spice / spice mix | Santa Maria | Homogenise d | Vortex mixing in buffer | 1.13 | 10.87 |
| 32 | Pasta rossa spice mix (garlic, tomato, and pepper) | Spice mix | Spice / spice mix | Santa Maria | Homogenise d | Vortex mixing in buffer | 1.13 | 10.87 |
| 33 | Chicken and steak spice mix (onion, paprika, and garlic) | Spice mix | Spice / spice mix | Santa Maria | Homogenise d | Vortex mixing in buffer | 1.13 | 10.84 |
| 34 | Barbeque spice mix | Spice mix | Spice / spice mix | Santa Maria | Homogenise d | Vortex mixing in buffer | 1.13 | 10.37 |
| 35 | Ras El Hanout spice mix (paprika, cayenne pepper, etc.) | Spice mix | Spice / spice mix | Santa Maria | Homogenise d | Vortex mixing in buffer | 1.13 | 9.70 |
| 36 | Tabasco green | Sauce | Processe d sauce | McIlhenny Company | Homogenise d | Vortex mixing in buffer | 1 | 9.52 |

| | | | | | | | | |
|---|---|---|---|---|---|---|---|---|
| | pepper sauce | | | | | | | |
| 37 | BBQ marinade | Spice mix | Spice / spice mix | Santa Maria | Homogenised | Vortex mixing in buffer | 12.5 | 8.96 |
| 38 | Paprika powder, smoked | Spice | Spice / spice mix | Santa Maria | Homogenised | Vortex mixing in buffer | 1.13 | 8.84 |
| 39 | Pasta sauce | Sauce | Processed sauce | Hello Fresh | Homogenised | Vortex mixing in buffer | 150 | 8.30 |
| 40 | Vegetable | Boullion cube | Spice / spice mix | Knorr | Homogenised | Vortex mixing in buffer | 5 | 8.08 |
| 41 | Tabasco | Pre-made sauce | Processed sauce | McIlhenny Company | Homogenised | Vortex mixing in buffer | 1 | 7.97 |
| 42 | Hot chili | Pre-made sauce | Processed sauce | Frank's | Homogenised | Vortex mixing in buffer | 15 | 7.80 |
| 43 | Instant noodle spice, spicy flavour | Spice mix | Spice / spice mix | Shamyang Ramen | Homogenised | Vortex mixing in buffer | 2.7 | 7.63 |
| 44 | Chili flakes | Spice | Spice / spice mix | Santa Maria | Homogenised | Vortex mixing in buffer | 1.13 | 7.22 |
| 45 | Chili | Capsicum fruit | Capsicum friut | ICA | Homogenised | Liquid nitrogen and grinding with a mortar and pestle | 4.5 | 7.18 |
| 46 | Cayenne pepper | Spice | Spice / spice mix | Santa Maria | Homogenised | Vortex mixing in buffer | 1.13 | 6.67 |
| 47 | Hamburger dressing | Pre-made sauce | Processed sauce | Kavli | Homogenised | Vortex mixing in buffer | 20 | 5.65 |
| 48 | Garam Masala | Spice mix | Spice / spice mix | Santa Maria | Homogenised | Vortex mixing in buffer | 1.13 | 5.65 |
| 49 | Cayenne pepper sauce | Pre-made sauce | Processed sauce | Buillards | Homogenised | Vortex mixing in buffer | 15 | 5.55 |
| 50 | Chili | Pre-made sauce | Processed sauce | Heinz | Homogenised | Vortex mixing in buffer | 15 | 5.52 |
| 51 | Piri piri sauce | Pre-made sauce | Processed sauce | FU Piri Piri | Homogenised | Vortex mixing in buffer | 15 | 5.45 |
| 52 | Swedish Falu sausage | Processed food | Processed food | Scan | Homogenised | Vortex mixing in buffer | < LOB. not estimated | < LOB |
| 53 | Oats | Negative control | Negative control | AXA | Homogenised | Liquid nitrogen and grinding with a mortar and pestle | < LOB. not estimated | < LOB |
| 54 | Spitzpaprika | Capsicum fruit | Capsicum friut | ICA | Homogenised | Liquid nitrogen and grinding with a mortar and pestle | < LOB. not estimated | < LOB |
| 55 | Sriracha | Pre-made sauce | Processed sauce | ICA | Homogenised | Vortex mixing in buffer | < LOB. not estimated | < LOB |
| 56 | Ketchup, containing Cayenne pepper | Pre-made sauce | Processed sauce | Felix | Homogenised | Vortex mixing in buffer | < LOB. not estimated | < LOB |
| 57 | Chili, home-made | Home-made sauce | Home-made food | Home-made | Homogenised | Vortex mixing in buffer | < LOB. not estimated | < LOB |
| 58 | Sweet chili | Pre-made sauce | Processed sauce | Santa Maria | Homogenised | Vortex mixing in buffer | < LOB. not estimated | < LOB |

| | | | | | | | | |
|---|---|---|---|---|---|---|---|---|
| 59 | Sweet and spicy BBQ | Pre-made sauce | Processe d sauce | Sweet Baby Ray's | Homogenise d | Vortex mixing in buffer | < LOB. not estimate d | < LOB |
| 60 | Asian style BBQ | Pre-made sauce | Processe d sauce | Santa Maria | Homogenise d | Vortex mixing in buffer | < LOB. not estimate d | < LOB |

***Supplementary Table 3.*** *Tested variants of Model 2*

| Model variant | Explanatory variables included | Akaike Information Criterion | Parameter coefficient estimates (associated *p*-values) |
|---|---|---|---|
| Model 2, variant 1 (final model) | $Log_{10}$ residents | -218 | 1.12 (< 0.001) |
| Model 2, variant 2 | $Log_{10}$ residents and socioeconomic index | -208 | 1.12 (< 0.001) and -0.000028 (0.99) |
| Model 2, variant 3 | $Log_{10}$ residents and $log_{10}$ proportion of foreign born residents | -215 | 1.08 (< 0.001) and 0.13 (0.32) |
| Model 2, variant 4 | $Log_{10}$ residents and longitude | -207 | 1.12 (< 0.001) and 0.000403 (0.94) |
| Model 2, variant 5 | $Log_{10}$ residents and latitude | -207 | 1.12 (< 0.001) and 0.00019 (0.99) |

***Supplementary Table 4.*** *Comparison of two extraction methods for three different food samples. Each sample was extracted in triplicate using both extraction methods, and the concentrations reported represent the average of these three extractions*

| Extraction kit | Tabasco, $log_{10}$ (gc/g) | Vegetable broth, $log_{10}$ (gc/g) | Chili powder, $log_{10}$ (gc/g) |
|---|---|---|---|
| Plant kit (Maxwell® RSC Plant RNA Kit) | 7.78 | 8.08 | 4.19 |
| Wastewater kit (Maxwell® RSC Enviro TNA kit, Promega) | 7.72 | 7.93 | 3.71 |
